# Current gaps in downstream applications of fMRI foundation models: lessons learned from BrainLM & BrainHarmonix on Prevent-AD

**DOI:** 10.64898/2026.10.22.26366227

**Authors:** Hao-Ting Wang, Alexa Pichet Binette, Pravish Sainath, Nicolas Farrugia, Sylvia Villeneuve, Lune Bellec

## Abstract

Foundation models have driven major advancements in the language and vision domains, as seen in the popularity of Artificial Intelligence (AI) tools, and a growing number of studies are bringing the approach to human brain imaging. Whether these models are ready for independent researchers to use on clinical datasets has yet to be examined. We evaluated two recent fMRI foundation models, BrainLM and BrainHarmonix, on PreventAD, a dataset on adults with a family history of Alzheimer’s disease. We compared features from frozen and fine-tuned models against two baselines: functional connectivity and a dummy classifier that always predicts the most common class. The sample includes 343 subjects who passed quality control, with a subset of 218 who had amyloid PET imaging. We tested four prediction targets: sex, age, progression to mild cognitive impairment (MCI), and β-amyloid (Aβ) burden. Both models required substantial changes to the released code. BrainLM’s own data preparation workflow did not produce stable fine-tuning, which we achieved only after switching to a standard workflow implemented with the library Nilearn. BrainHarmonix’s documentation did not specify how to normalize new data, so we resolved this by testing different alternatives. For sex, functional connectivity outperformed all foundation model features. For age group, the best foundation model feature, taken from BrainHarmonix’s structural (T1-weighted) encoder, slightly exceeded functional connectivity, but the difference was not significant. For MCI and Aβ status, no feature exceeded the dummy classifier in accuracy, and the area under the curve (AUC) stayed near chance for all features. For both clinical targets, the most precise feature came from fine-tuned BrainLM, meaning a larger share of its positive predictions were correct than for functional connectivity, but neither difference reached statistical significance. These results suggested that the strong transfer to new tasks seen in large language models cannot yet be assumed for fMRI foundation models, and that both models required code adaptation that would preclude their streamlined adoption by independent groups. We recommend that future studies provide standardized data workflows and always report functional connectivity and dummy classifier baselines. For clinical targets with strong class imbalance, studies should report several metrics, including precision.

## Introduction

Foundation models are a critical recent research avenue for capitalizing on large-scale, open, and heterogeneous data to learn general representations that are transferable across samples (Bommasani et al., 2021). These models are pre-trained in a self-supervised regime, which means they do not rely on task-specific labels, such as age or clinical diagnosis. Instead, foundation models learn the distribution of the data itself, typically through reconstructing parts of the data which have been masked, which is key to enabling broad pooling of data sources that do not carry homogeneous labels. The representations they learn can be reused with minimal, if any, modifications for downstream tasks, such as making diagnostic and prognostic decisions in small, independent datasets. This framework was key to many recent advances in Artificial Intelligence (AI), notably Large Language Models (LLMs) such as ChatGPT (Brown et al., 2020). A large number of recent works aim to adapt this idea in the field of neuroimaging, for instance, training a foundation model on a large set of unlabeled brain data, such as the UK Biobank (Sudlow et al., 2015), and then using the representations of brain data learned by the model in order to diagnose neurological or psychiatric conditions such as Attention Deficit Hyperactivity Disorder (ADHD) or Autism in separate data samples. This line of research is still in its infancy for functional magnetic resonance imaging (fMRI), and it remains unclear whether the models can be readily applied by practitioners to real-world clinical samples of modest sample size. In this work, we aim to evaluate two recent fMRI foundation models on challenging downstream diagnosis and prognosis tasks in preclinical Alzheimer’s disease (AD).

FMRI foundation models emerged in 2023 (Zhou et al., 2025), including BrainLM (Caro et al., 2023), Brain-JEPA (Dong et al., 2024), and BrainHarmonix (Dong et al., 2025), the latter two from the same research group, with each successive model reporting improvements over prior work. These models were trained on large population-level datasets and were evaluated on open clinical datasets. This experimental setup assumes that information from the foundation model can be meaningfully transferred to downstream datasets. We argue that rigorous evaluation of the models is needed before we can assume the robust generalization capabilities seen in LLMs are also a property of fMRI foundation models.

Several gaps remain in the assessment of current fMRI foundation models. Firstly, other research groups seeking to apply these models to their own datasets face an untested generalization problem in terms of practical implementation: it is unclear whether models trained and evaluated within a single lab’s pipeline will perform reliably on new datasets, and whether the documentation practices are sufficient for a new group of researchers to implement the workflow for feature extraction, fine-tuning, and downstream applications. Furthermore, functional connectivity, the current standard feature for fMRI-based trait prediction (Dadi et al., 2019; Finn et al., 2015; Khosla et al., 2019), is absent from existing foundation model benchmarks, making it impossible to assess whether foundation models improve on well-established baselines.

We propose to address these two gaps on a challenging benchmark with data collected on older adults at risk of preclinical AD participants from the PREVENT-AD cohort. Late-onset AD is the most prevalent age-related neurodegenerative condition and a rising public health concern in an ageing Canadian population. AD is a complex, multifactorial pathology (Badhwar et al., 2020) that manifests along a continuum of cognitive symptoms from cognitively normal to prodromal mild cognitive impairment (MCI) to dementia (Alzheimer’s disease dementia, ADD). AD pathology starts to deposit in the brain decades before cognitive symptoms appear, in the so-called preclinical phase of the disease (Sperling et al., 2011), which is a critical period during which to intervene to prevent or delay the disease. We used the PREVENT-AD cohort to test whether foundation models could help identify reliable predictive features in this preclinical phase of the disease, as well as whether it could predict individuals who will develop cognitive decline.

This work aims to achieve three objectives. Firstly, we identify the challenges an independent group of researchers would face when deploying foundation models on a new dataset. We expect that, even with the source code and model weights available, we will still need to adapt the code due to the lack of standard libraries for data processing. Secondly, we compare the foundation models with a common baseline in the fMRI machine learning literature. We hypothesize that foundation model features would perform comparably, rather than decisively outperform, established baselines, as observed in LLM benchmarks (Brown et al., 2020). Lastly, we examine whether foundation models improve performance in downstream applications and, if so, which specific models and types of tasks they perform well on. We speculate that foundation models would have higher and more robust performance on downstream tasks than baseline models, especially in the presence of low signal and smaller sample sizes.

Here, we evaluate BrainLM, one of the first fMRI foundation models trained on resting-state data, and BrainHarmonix, the most recent state-of-the-art model from the same lab that produced Brain-JEPA, which BrainHarmonix supersedes on comparable benchmarks, on the Pre-symptomatic Evaluation of Experimental or Novel Treatments for Alzheimer’s Disease (Prevent-AD) dataset (Villeneuve et al., 2025), an independent cohort not used in the training or evaluation of either model, to test independent generalization directly. We include functional connectivity as a baseline feature and assess downstream prediction performance for sex, age, MCI conversion status, and amyloid-beta (Aβ) burden, covering both demographic targets and outcomes relevant to stratifying individuals along the AD continuum. Together, these evaluations provide an external benchmark of fMRI foundation models from a user perspective, establishing whether current models are ready for adoption by independent research groups working on neurodegeneration and cognitive decline.

## Methods

### Foundation Models

We selected two fMRI foundation models and downloaded pretrained model checkpoints from their respective sources, as described in the sections below. The detailed application of the models will be described in the Evaluation Framework - Feature extraction section.

**BrainLM** (Caro et al., 2023) is an early fMRI foundation model trained on resting-state fMRI. Its architecture is based on a transformer-masked autoencoder (similar to ViT-MAE for images; He et al., 2021) to learn the spatiotemporal dynamics of fMRI. BrainLM is pretrained on the UK Biobank using a masked prediction objective, in which a percentage of the input time-series data is randomly masked, and the visible (unmasked) inputs are used to reconstruct the original signals of the masked parcels. The model produces latent representations as classification tokens (Stylised as CLS tokens to remain consistent with the literature; Devlin et al., 2018) and embeddings, which are then used for downstream tasks. The pre-trained model was available in two sizes: 111M and 650M parameters. We downloaded the model checkpoints from the public Hugging Face repository and followed the instructions in the GitHub repository to extract features. The training data were fMRI time series extracted using the A424 atlas (Nemati et al., 2020). For extraction procedures, see Section Dataset - Post-fMRIPrep denoising and time series extraction. We evaluated only the larger pretrained model (650M) because the original paper demonstrated that it performs better. The model checkpoints were downloaded from HuggingFace (https://huggingface.co/vandijklab/brainlm).

**BrainHarmonix** (Dong et al., 2025) is a multimodal brain foundation model. It unifies structural images and functional dynamics into compact 1D token representations. BrainHarmonix is grounded in two foundational neuroscience principles: (1) structure complements function: structural and functional modalities offer distinct yet synergistic insights into brain organization; (2) function follows structure: brain functional dynamics are shaped by cortical morphology. The modular pretraining process involves single-modality training with geometric pre-alignment to existing functional gradients (Margulies et al., 2016), followed by modality fusion through shared brain hub tokens. Notably, the dynamics encoder handles fMRI time series with heterogeneous repetition times (TRs), addressing a major limitation in existing models. BrainHarmonix is also the first to deeply compress high-dimensional neuroimaging signals into unified, continuous 1D tokens, forming a compact latent space of the human brain. The training data were fMRI time series extracted using the Schaefer 400 atlas (Schaefer et al., 2018). For extraction procedures, see Section Dataset - Post-fMRIPrep denoising and time series extraction. The model checkpoints were downloaded from Google Drive hosted by the authors (https://drive.google.com/drive/folders/12MkUAOcegU60YVlK8u8_Owmgk4eQVheB).

### Downstream Dataset

The present study utilized data from the Prevent-AD (Pre-symptomatic Evaluation of Experimental or Novel Treatments for Alzheimer’s Disease) cohort. The Prevent-AD cohort is composed of cognitively unimpaired individuals with a familial history of AD, which is a known substantial risk factor, and who are followed up every year. The cohort is created to study pre-symptomatic changes from a pathophysiological perspective, including biochemical changes and structural or functional alterations in the brain. The study used only openly available human data collected before the study began. The dataset is hosted and shared through the Prevent-AD LORIS platform, where we obtained data access (https://preventad.loris.ca).

We focus exclusively on the baseline session and resting-state functional Magnetic Resonance Imaging (rs-fMRI) run 1. Details of the sequence can be found in the original dataset description (Villeneuve et al., 2025). 381 subjects completed the baseline session, and 343 passed quality control (see section Resting-state fMRI preprocessing and quality control). The cohort showed a female bias, with 254 females and 89 males. Subjects’ ages at baseline ranged from 55 to 84 years (Mean Age (SD): 63.51 (5.09) years). A subsample of 218 subjects underwent PET imaging and was used for prediction tasks related to pathophysiology and the emergence of clinical symptoms. See Figure 1A for a summary of samples.

**Figure 1.**
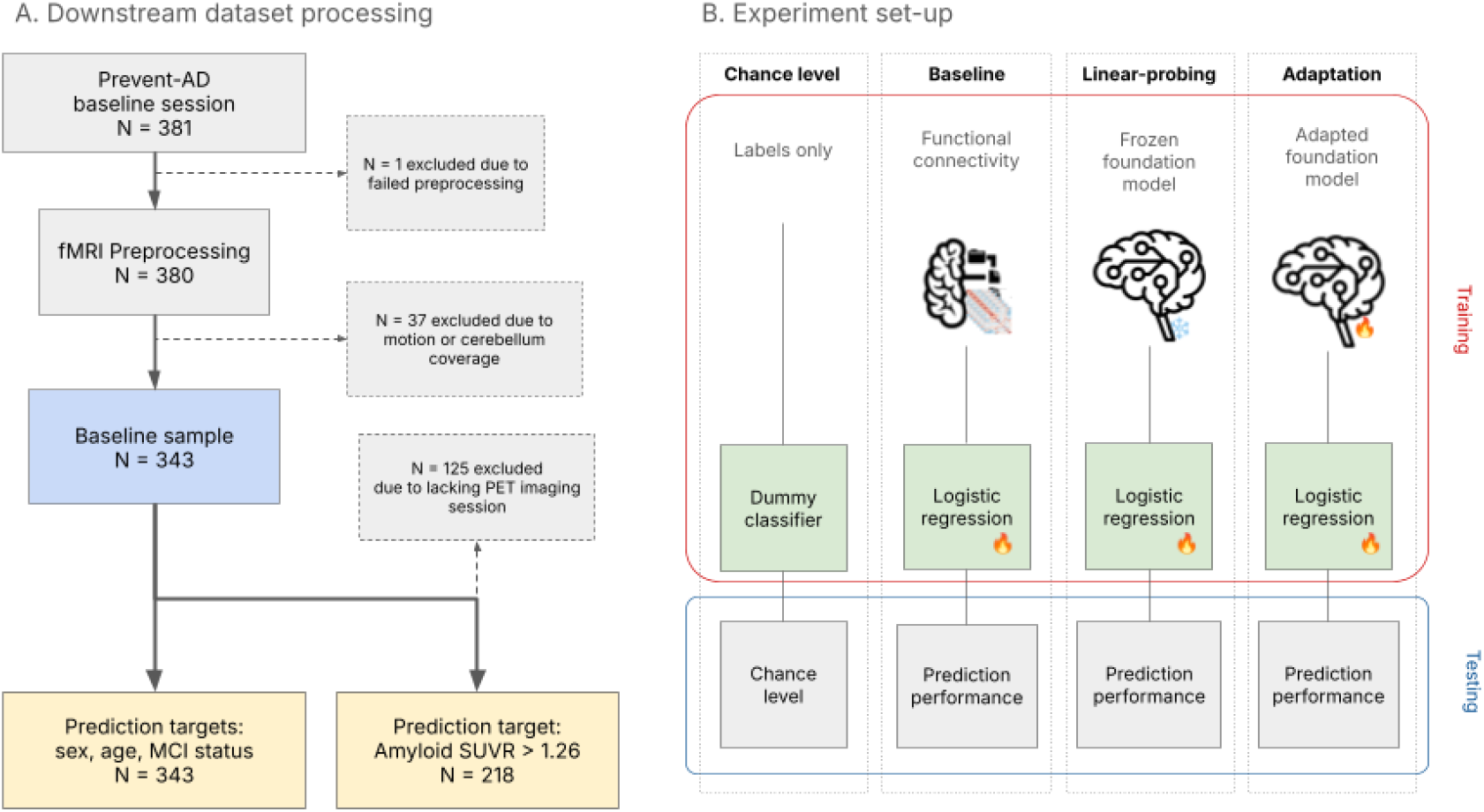
Summary of the sample selection process. (A) Sample selection process. The solid-line rectangular boxes represent datasets, and the dashed-line boxes detail the reasons for exclusion at different stages of quality control. The baseline sample indicated by the blue box passed all fMRI data quality control, and the yellow boxes indicate the sample and its respective prediction target. (B) Four approaches are compared to assess feature quality for downstream trait prediction: a dummy classifier serving as a chance-level reference, functional connectivity as a standard baseline, a frozen foundation model with fixed pretrained weights, and a adapted foundation model with weights updated on the target dataset with a self-supervised learning objective. All approaches use logistic regression as the downstream classifier.

#### Downstream prediction targets

We selected basic demographic information (sex and age) and MCI-specific traits as prediction targets. Sex and age are shared phenotypic information associated with all MRI data. They can serve as a reference for assessing whether the extracted features contain detectable signals when the sample size is maximized. The AD-specific traits are targets of clinical interest, with future MCI status as a prognosis target for AD and Aβ status as a target of preclinical AD.

All information comes from the Prevent AD dataset. Details are as follows:

- Sex (N=343; full experiment sample): information collected for the imaging session used in the analysis (i.e., the baseline session). There are two classes (Male / Female).
- Age (N=343; full experiment sample): information collected for the imaging session used in the analysis (i.e., the baseline session). We used the median as the threshold to create two balanced classes as the prediction target due to the narrow age range (Mean Age (SD): 63.51 (5.09) years).
- MCI status (N=343; full experiment sample; N_positive_ = 59): whether a subject develops MCI in the future.
- Aβ status: The measure was quantified through Aβ Standardized uptake value ratio (SUVR), a PET-based biomarker (N=218; subsample; N_positive_ = 76); a quantitative measure of Aβ accumulation. SUVR were derived using cerebellum grey matter as the reference region and average neocortical SUVR was calculated. We used 1.26 as the cutoff threshold to create two classes, as per done by the PREVENT-AD group (Jack et al., 2017; Villeneuve et al., 2025).

#### Resting state fMRI preprocessing and quality control

We preprocessed fMRI data using fMRIPrepLTS (Esteban et al., 2019; RRID:SCR_016216; version 20.2.8 LTS). One subject was excluded because it failed to run through fMRIPrep. We have two quality-control criteria: motion and cerebellar coverage. For motion, scans were excluded if the percentage of volumes marked as high motion (framewise displacement >0.5 mm) exceeded 50% of the total run duration. Because BrainLM uses an atlas that covers the cerebellum, we included only scans with cerebellar coverage greater than 75% for further analysis to avoid coverage-related bias, as more male subjects have incomplete cerebellar coverage. See Figure 1A for the graphical summary of the process.

#### Post-fMRIPrep denoising and time series extraction

For all experiments, whether or not foundation models are involved, we performed post-fMRIPrep denoising and extracted time series from the fMRIPrep output.

We adopted two different time-series extraction workflows: one shipped with BrainLM and the other is giga-connectome (Wang et al., 2025), a standardized workflow built on Nilearn. BrainLM workflow handled only time-series extraction and provided multiple normalization options, while giga-connectome was designed to denoise, extract, and normalize fMRI data preprocessed with fMRIPrep. BrainLM’s supplied normalization workflow includes hard-coded scaling values and the replacement of missing values with 0. We decided to compare whether a more standardized workflow that does the same type of scaling in principle and centres the data to 0 makes a difference. To ensure the differences arise solely from the normalization methods, we denoised the data in the same manner for both workflows, using six motion parameters, the white matter and CSF tissue signals, and the global signal. The reported normalization method in the original paper was a robust scalar. In the giga-connectome workflow, we used the scikit-learn implementation, and in the BrainLM workflow, we used “Subtract_Mean_Divide_Global_STD_Normalized_Recording”. A summary and comparison of the details are listed in Table 1.

**Table 1.** Timeseries extraction workflow.

|  | **BrainLM** |  |  |  | **BrainHarmonix** |  |
|---|---|---|---|---|---|---|
| Preprocessing | fMRIPrep<br>Functional MRI "Prep" |  |  |  |  |  |
| fMRI voxel level post processing | Denosing: high-pass filter, 6 motion parameters + WM + CSF + global signal |  |  |  |  |  |
|  | Within subject normalization:<br>z-score |  | No normalization |  | Within subject normalization: z-score | No normalization |
| Atlas | A424 |  |  |  | Schaefer 400 |  |
| Time series extraction | BrainLM workflow | Nilearn workflow | BrainLM workflow | Nilearn workflow | Nilearn workflow | Nilearn workflow |
| Global normalization** | BrainLM workflow | Robust scalar | BrainLM workflow | Robust scalar | Robust scalar | Robust scalar |

Workflow matching neuroimaging research standard (giga-connectome workflow)
Closest to original workflow based on released code
Outputs for generating benchmark baseline
\*\*Only applied for foundation model training / adaptation /  fine-tuning

### Evaluation Framework

We evaluated the performance of the foundation models by their ability to classify targets on a new dataset, as detailed in the “Downstream Dataset” section. The general process of the evaluation framework is illustrated in Figure 1B. We performed four sets of experiments: (1) <u>Chance level</u>: a dummy classifier predicting the majority class, establishing the chance level and serving as the lower bound of performance; (2) <u>Baseline</u>: classification using functional connectome and timeseries served as a non-foundation model comparison baseline; (3) <u>Linear-probing</u>: classification using frozen foundation model representations; and (4) <u>Adaptation</u>: classification on adaptedfoundation model representations.

Here, we first report the common procedures before describing each experiment in detail. We used regularised logistic regression (the default scikit-learn implementation) as the downstream classifier in all non-chance experiments (experiments 2, 3, and 4). We did not include other classifiers to isolate feature quality from classifier choice. In each experiment, we performed twenty 80%-20% train-test shuffle splits, stratified by the prediction target to preserve class proportions for cross-validation. We summarised classifier performance on the test sets across splits.

#### Baseline: timeseries and functional connectome

Time series were included as a baseline feature because they were used in the downstream evaluation of BrainLM. In the BrainLM experiment, the top 5 principal components of the time series were used as baselines for comparison; we included the same components in the current experiment, with one change. Instead of using the top 5 components, we used the top 75 components. The neuroimaging literature shows that the top decompositions likely reflect the principal organization of the brain signal (Margulies et al., 2016). By extracting more components, we hoped to capture individual differences better.

We also included the functional connectome as the baseline. In the fMRI-based trait prediction framework, the functional connectome is a commonly used default feature (Dadi et al., 2019). A functional connectome is a set of connections (i.e., functional connectivity) representing interactions in brain activity between regions. In this benchmark, we used time series extracted with the Schaefer 400 atlas (Schaefer et al., 2018) and A424 (Nemati et al., 2020). The functional connectivity was calculated as Pearson’s Correlation between regional timeseries implemented via nilearn.connectome.ConnectivityMeasure. The 400 regions create 79,800 pairs of functional connectivities ((400 * 400 - 400) / 2).

#### Linear-probing: Feature extraction from frozen foundation models

In this experiment, pretrained weights were held fixed and representations (“frozen”) were extracted without any parameter updates. For BrainLM, we extracted the attention head of the CLS Token, the CLS embedding (unprocessed, mean-pooled, and max-pooled). For BrainHarmonix, we extracted four feature sets, corresponding to different stages of its modular architecture. The *fMRI (mean-pooling)* and *T1 (mean-pooling)* were the mean-pooled latent features from the single-modality functional and structural encoders, respectively, prior to modality fusion. The *Harmonizer (CLS)* and *Harmonizer (latent tokens mean-pooling)* features are extracted after fusion through the shared brain hub tokens: the Harmonizer is a transformer module, and the CLS feature corresponds to its classification token summarising the fused representation, while the latent feature is the mean-pooled output across the fused token sequence.

#### Adaptation: Foundation model updated with new data

For the adaptation experiment, we updated the model using the same self-supervised masked reconstruction objective as in the original pretraining procedure. We used the models adapted on the training set to extract features for downstream evaluations, and the procedures were identical to those described in the linear-probing experiment section above. We deliberately refrained from performing an extensive hyperparameter search due to the number of conditions introduced by the normalization options (see Table 1) and to provide a realistic, limited-tuning deployment representative of their likely use by researchers outside for the original author’s teams.

Hyperparameter values for adaptation were adjusted empirically based on convergence behaviour observed during preliminary runs, anchored around PyTorch’s AdamW defaults (learning rate 1e-3, weight decay 0.01; Loshchilov & Hutter, 2017). Both BrainLM and BrainHarmonix were fine-tuned with the AdamW optimizer, a learning rate of 1e-4, within the range conventionally used for fine-tuning pretrained transformer and vision transformer models rather than training from scratch (Devlin et al., 2018; He et al., 2021), a weight decay of 0.01, and a batch size of 4. We fine-tuned both models for up to 50 epochs with early stopping, halting training when validation loss failed to improve for 5 consecutive epochs (patience = 5), and applied this independently to each train/test split (Prechelt, 2012). We retained the checkpoint with the lowest validation loss for each split and used it for downstream feature extraction.

#### Metrics

We treated all prediction targets as binary classification problems. We reported accuracy, Area Under the Curve (AUC), F1 score, and precision for each target and condition. AUC accounts for class imbalance by averaging recall across classes, and is reported alongside precision to provide a complete picture of model performance on skewed clinical samples (Clarke et al., 2025). We summarised performance as the mean and 95% confidence interval across the stratified shuffle splits. For comparisons of metrics with the functional connectivity baseline, we used an unequal-variance t-test. For comparison with chance level, we used a one-sample t-test.

## Results

### Challenges of deploying foundation models on new datasets as an independent research group

When reviewing and adopting the BrainLM and BrainHarmonix codebases, we found a few issues that required some customizations.

#### Issues with signal normalization

The consistency of normalization practices across the datasets used by both models was unclear; hence, we implemented multiple options (see Table 1). We used the best denoising strategy for continuous time series based on a previous benchmark (Wang et al., 2024). After nuisance regression, we generated two versions of the voxel-level normalized data: with or without grand-mean scaling. The reason was that BrainLM explicitly performed grand-mean scaling in their preprocessing workflow to centre the signal around 0. It was unclear if data used in BrainHarmonix were scaled in the same manner; hence, we performed both.

#### Checkpoint sharing

For BrainLM, the model was available for download via the Hugging Face library. Since the model could be loaded via the standard Hugging Face library, it was straightforward to load it from the source code and the paper, which describes its architecture. For BrainHarmonix, the model was downloaded through Google Drive rather than a more secure open repository.

#### Source code quality

For BrainLM, the code implemented the full time-series extraction procedure without using common libraries such as Nilearn, and it relied on all fMRI files being sampled in the same standard space and masked with whole-brain masks. The robust scalar, a type of normalization procedure, was implemented by the authors themselves, and it was incorrect. Several issues are present on the issue page yet remain unaddressed. For BrainHarmonix, the source code was not installable despite being packaged. Fortunately, we found a fixed fork maintained by MedARC-AI that fixed the installation issue. Yet, some manual fixes were needed, so we used a customized fork for this project (https://github.com/htwangtw/Brain-Harmony/tree/fix/flash-attn-version-shadowing).

#### Self-supervised learning adapation learning curves and model exclusion

We implemented both models and found some fMRI data-processing options led to problematic learning curves (Figure 2), hence were removed from the subsequent report. For BrainHarmonix, the learning curves using data with no global normalization (Figure 2b) were noisy, with a few cross-validation splits failing to converge within 50 epochs. BrainHarmonix with global voxelwise normalization (Figure 2a) showed stable learning curves. For BrainLM, the only stable learning curve uses the giga-connectome workflow with no global voxelwise normalization, yet the magnitude of loss change was small (Figure 2f). Note that all learning curves with BrainLM provided preprocessing showed unusual patterns (Figure 2c and Figure 2d). In the following report, we will only report results with stable learning curves (Figure 2a and Figure 2f).

**Figure 2.**
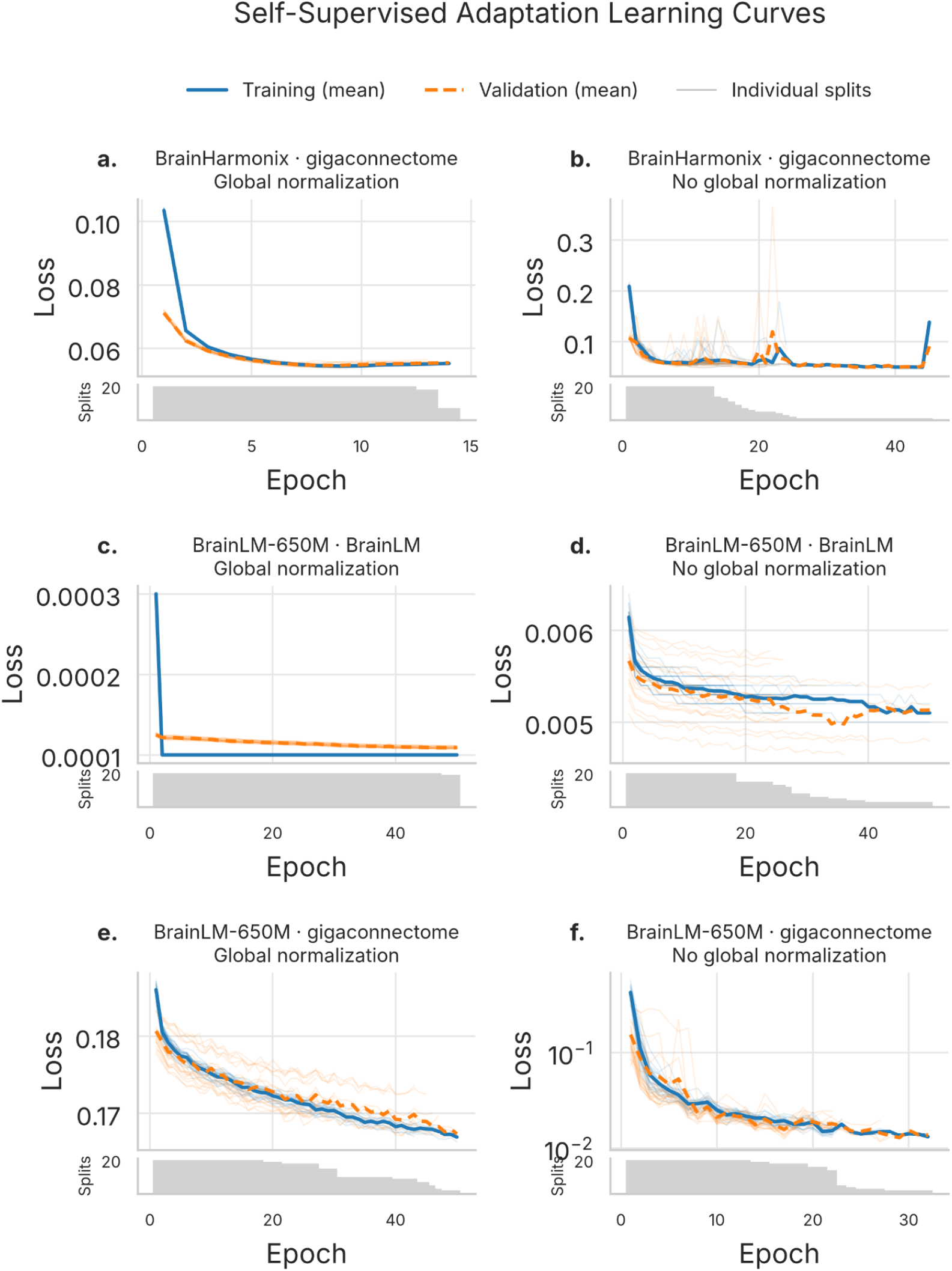
Self-supervised adaptation learning curves. Columns correspond to global voxelwise normalization (left) versus no normalization (right). Each panel shows training (solid blue) and validation (dashed orange) loss as a function of epoch, averaged across 20 cross-validation splits (bold lines), with individual splits shown as faint lines. The bottom-right panel uses a logarithmic y-axis, where loss spans more than one order of magnitude. The grey strip beneath each panel shows the number of splits still contributing at each epoch; because training used early stopping, later epochs are supported by progressively fewer splits, so interpret mean curves in the right-hand tail of each panel with correspondingly reduced confidence.

### Functional connectivity baseline: establishing the reference feature

For each prediction target, we established two references for comparison: a scikit-learn dummy classifier and functional connectivity. The dummy classifier accounted only for the prediction target; its accuracy score equalled the proportion of the majority class in the target, so we did not report confidence intervals for it. Functional connectivity is a common feature in the fMRI machine learning literature and serves as a performance baseline. In Figure 3, we also added time series as a comparison, as it was featured in the BrainLM benchmark to demonstrate it performs at chance level in both accuracy and AUC; thus, it shouldn’t serve as a baseline representing fMRI-derived features. We report the dummy classifier and functional connectivity metrics in Table 2.

**Figure 3.**
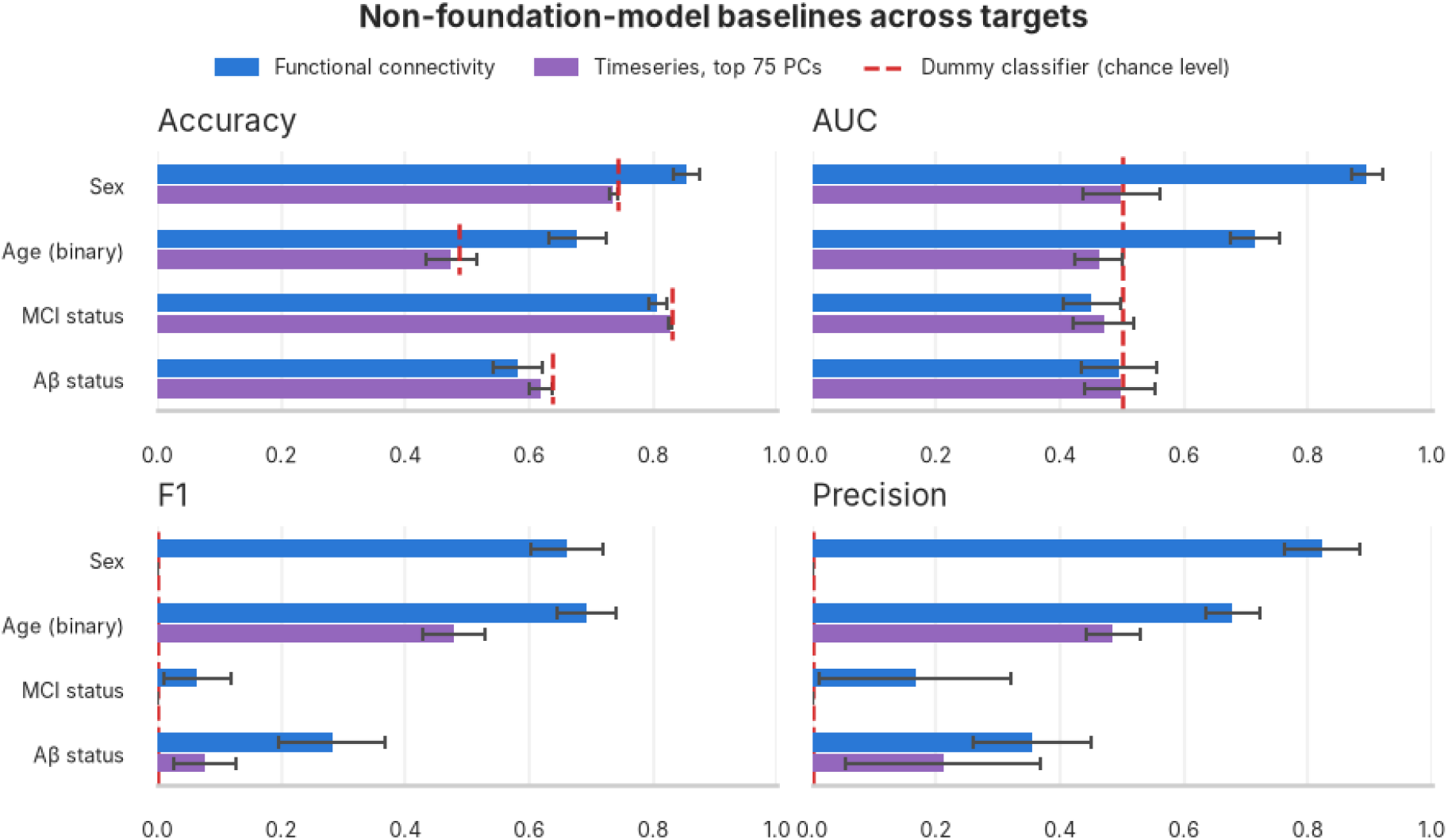
Non-foundation model baselines across targets. We showed baselines using functional connectivity, a common feature in fMRI machine learning literature, and time series, the baseline feature used in BrainLM, as features, and chancel level performance denoted as dash lines. Time series performed close to chance in accuracy. Sex and age group predictions are the only target outperforming chance level baseline.

**Table 2.** Baseline performance.

| **Target** | **Feature** | **Accuracy** | **AUC** | **F1** | **Precision** |
|---|---|---|---|---|---|
| Sex | Functional connectivity | 0.854<br>[0.833 0.875] | 0.895<br>[0.871 0.919] | 0.662<br>[0.603 0.720] | 0.823<br>[0.761 0.884] |
|  | Dummy classifier | 0.743<br>[- -] | 0.500<br>[- -] | 0.000<br>[- -] | 0.000<br>[- -] |
| Age<br>(median split) | Functional connectivity | 0.677<br>[0.631 0.723] | 0.715<br>[0.675 0.755] | 0.693<br>[0.646 0.739] | 0.679<br>[0.635 0.722] |
|  | Dummy classifier | 0.486<br>[- -] | 0.500<br>[- -] | 0.000<br>[- -] | 0.000<br>[- -] |
| MCI Status | Functional connectivity | 0.807<br>[0.793 0.821] | 0.451<br>[0.404 0.498] | 0.065<br>[0.011 0.119] | 0.167<br>[0.013 0.321] |
|  | Dummy classifier | 0.829<br>[- -] | 0.500<br>[- -] | 0.000<br>[- -] | 0.000<br>[- -] |
| Aβ status | Functional connectivity | 0.582<br>[0.542 0.621] | 0.495<br>[0.435 0.555] | 0.282<br>[0.196 0.367] | 0.355<br>[0.260 0.449] |
|  | Dummy classifier | 0.636<br>[- -] | 0.500<br>[- -] | 0.000<br>[- -] | 0.000<br>[- -] |

For sex and age group prediction, functional connectivity achieves performance above the majority-class baseline established by the Dummy classifier. For MCI and Aβ status, functional connectivity accuracy falls below that of the dummy classifier, reflecting the class imbalance inherent in these targets, and the AUC remains near 0.5, indicating limited discriminative ability. Average precision scores for both targets were nonzero, but the lower bound of the 95% confidence intervals is close to zero for MCI status.

In the following sections, we report only the conditions that outperform the dummy classifier baseline and use precision scores to screen potentially relevant methods, even though the functional connectivity baseline’s precision may not be significant in some experiments. Our aim was to screen potentially relevant foundation model features, even though the baseline’s precision may not be significant in some tests.

### Sex and age group prediction validates that the foundation model features carry meaningful but weak signals

For sex and age group prediction, many features extracted from foundation models performed above the dummy classifier; see Figure 4. The best-performing foundation model feature for both sex and age group prediction was latent weights of BrainHarmonix’s frozen T1 encoder (sex: average accuracy = 0.841 CI 95% [0.823 0.859]; age: average accuracy = 0.705 CI 95% [0.673 0.737]). For age group prediction, average accuracy was higher than the functional connectivity baseline, but the difference was not significant (*t* (34.0) = 1.05, *p* = 0.15). Overall, BrainHarmonix features performed better than BrainLM.

**Figure 4.**
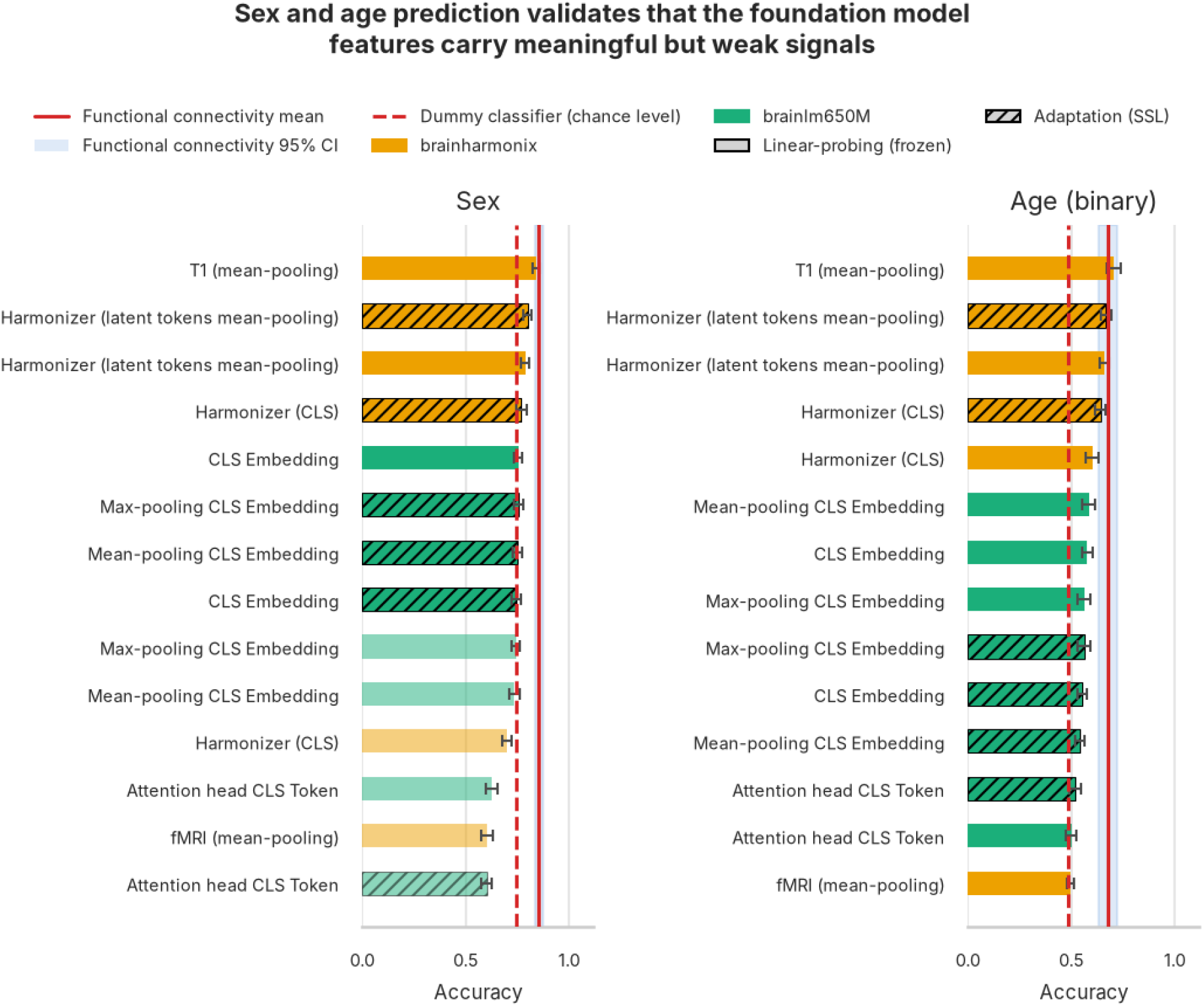
Sex and age group prediction. Classification accuracy for sex (left) and a binarized age variable (right) from two foundation models, BrainHarmonix (orange) and BrainLM (green), using either frozen or adapted (hatched) representations, ranked by accuracy within each panel. The solid red line and shaded band mark the functional-connectivity baseline’s mean and 95% CI; the dashed red line marks the dummy classifier’s chance level. Bars whose mean accuracy falls below chance are shown at reduced opacity. Bars are the mean across cross-validation splits; error bars are the 95% CI.

### MCI status and Aβ status prediction reveal a precision advantage for foundation-model-derived features

No foundation-model-derived feature outperformed the dummy classifier in accuracy, and all AUCs indicated near-chance performance. Several features achieved prediction precision above the functional connectivity baseline but did not reach statistical significance. Figures 5 and 6 report results ranked by precision scores. The supplemental material includes full summary statistics.

**Figure 5.**
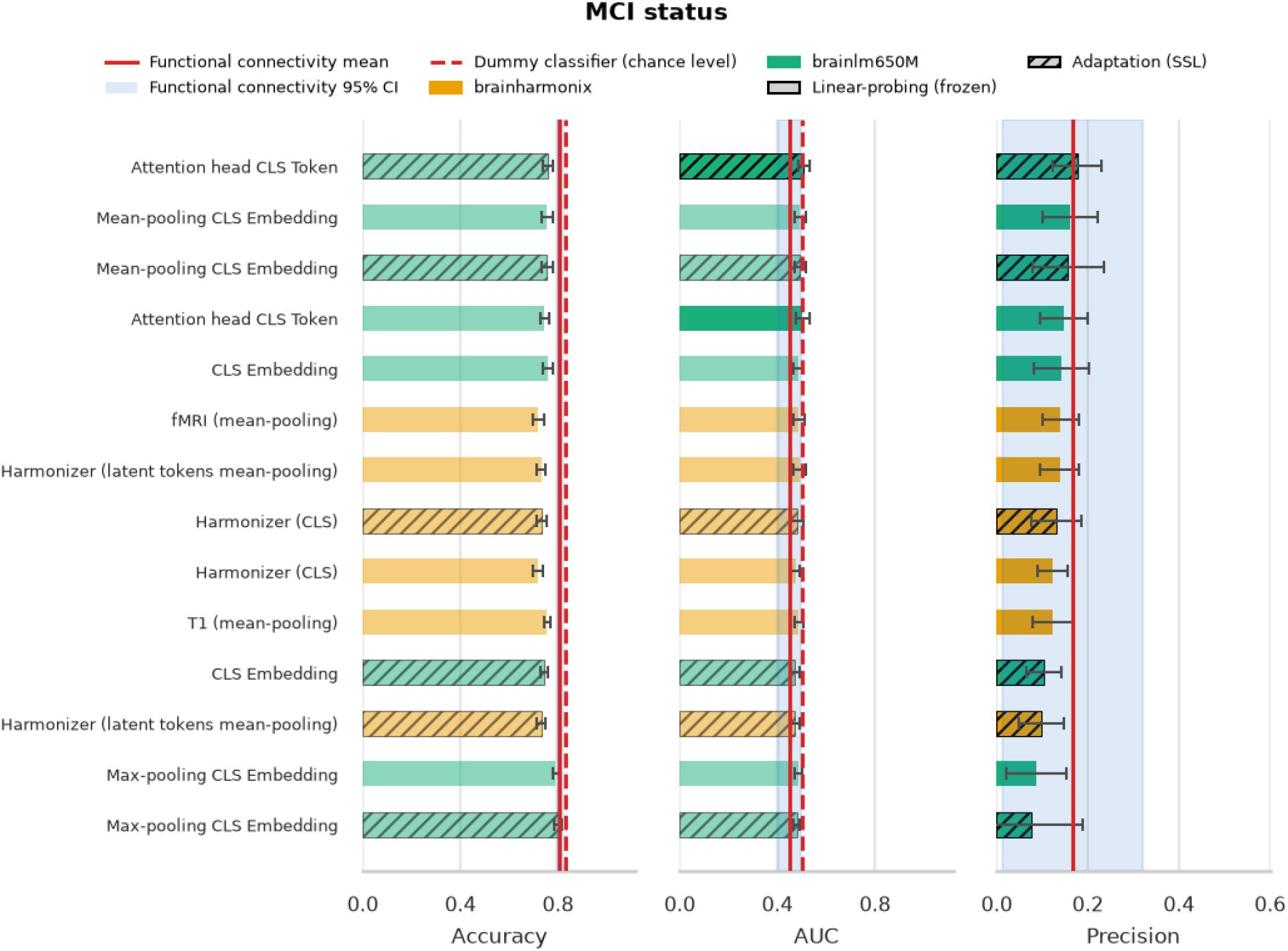
MCI status downstream. Accuracy, AUC and precision for classifying MCI status from BrainHarmonix (orange) and BrainLM (green) features, using frozen or adapted (hatched) representations. Rows are ranked by precision and are shared across panels. The solid red line and shaded band mark the functional-connectivity baseline’s mean and 95% CI. The dashed red line marks the dummy classifier’s chance level for accuracy and AUC; precision has no chance line because the dummy classifier never predicts the positive class. Bars whose mean falls below chance for that metric are shown at reduced opacity. Bars are means across cross-validation splits, and error bars are 95% CIs.

**Figure 6.**
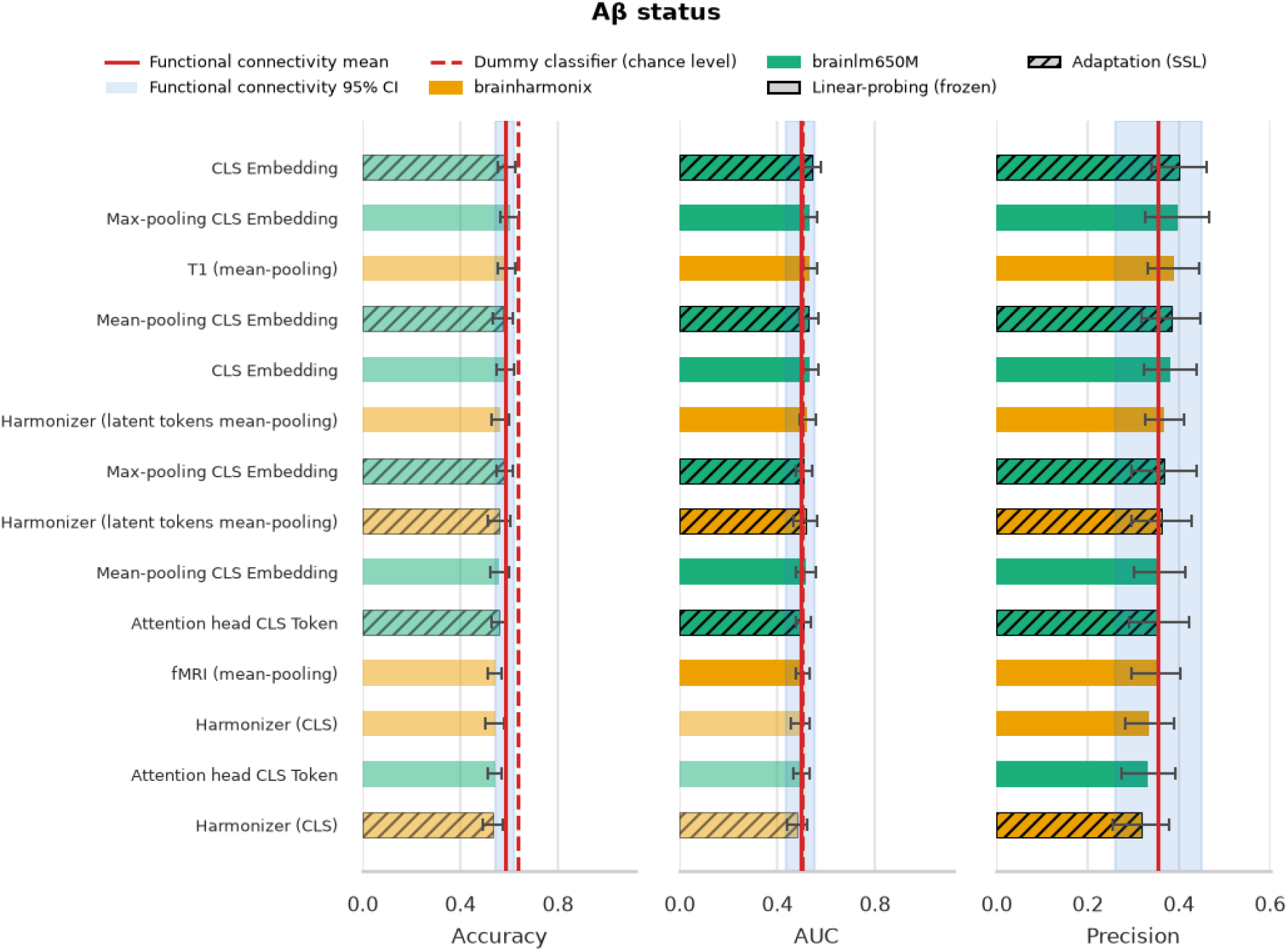
Aβ status downstream. Accuracy, AUC and precision for classifying Aβ status from BrainHarmonix (orange) and BrainLM (green) features, using frozen or adapted (hatched)representations. Rows are ranked by precision and are shared across panels. The solid red line and shaded band mark the functional-connectivity baseline’s mean and 95% CI. The dashed red line marks the dummy classifier’s chance level for accuracy and AUC; precision has no chance line because the dummy classifier never predicts the positive class. Bars whose mean falls below chance for that metric are shown at reduced opacity. Bars are means across cross-validation splits, and error bars are 95% CIs.

For MCI status (Figure 5), the fine-tuned BrainLM attention head CLS token achieved the highest precision (mean=0.176; 95% CI [0.122 0.231]) and exceeded the functional connectivity baseline (mean=0.167; 95% CI [0.013 0.321]) but did not reach statistical significance. For Aβ status (Figure 5), the fine-tuned BrainLM CLS embedding (mean=0.40; 95% CI [0.339 0.461]) was the top performer and outperformed the functional connectivity baseline (mean=0.355; 95% CI [0.260 0.449]) in precision but did not reach statistical significance.

## Discussion

Foundation models rapidly gained popularity in machine learning research following their success in LLMs. Several fMRI foundation models have recently been developed, but their performance when independent research groups apply them to new datasets and use them to predict clinical symptoms remains untested. Here, we benchmarked two such models, BrainLM and BrainHarmonix, against a functional connectivity baseline on the Prevent-AD dataset, testing independent generalization from a user perspective.

BrainLM could not be adapted using its source code timeseries extraction workflow, as training loss failed to converge; convergence was achieved only after adopting a standardized neuroimaging extraction pipeline. On sex prediction, functional connectivity outperformed all foundation-model-derived features, including adaptation variants. On age group prediction, one foundation-model-derived feature outperformed functional connectivity but did not reach statistical significance. For clinical targets, no feature outperformed the dummy classifier baseline in terms of accuracy, given the severe class imbalance in MCI conversion status and Aβ status. Foundation model features achieved higher precision than functional connectivity in some cases. Although the differences did not reach statistical significance, it is worth exploring further with improved adaptation and fine-tuning procedures and investigating the potentially clinically relevant signal capture by the model compared with the functional connectivity baseline.

### Independent application of foundation models was technically challenging

The code for both foundation models was released in public repositories, and the original papers detail the data-processing steps. In theory, these practices should enable independent research groups to adopt the pretrained models for new projects. In practice, however, we faced challenges adopting either workflow, showing that code and documentation do not guarantee straightforward reuse.

With BrainLM, we found the source code relied on multiple hardcoded scaling factors and a custom time-series extraction workflow built from scratch rather than the community-standard library Nilearn. This workflow was highly specific to the original training data and may omit steps needed to apply it to independent datasets. The complexity of this custom code made it difficult to isolate and troubleshoot, and it produced noisy learning curves during self-supervised learning update. The loss statistics changed in very small magnitudes, and this was likely the source of the degenerate learning curve reported in Figure 2c. The only working variation uses a feature extraction workflow closer to the fMRI research community standard.

BrainHarmonix’s codebase was more narrowly scoped to model training and downstream application, and the accompanying paper clearly detailed its data-processing steps. Even so, we struggled to determine the correct data normalization procedure when applying the model to our own dataset. We tested two plausible normalization approaches and found one variation with a more stable learning curve, though the ambiguity itself remains a documentation gap.

These cases suggested the current implementation of fMRI foundation models raises substantial technical challenges for user adoption. Standardized software is crucial for broad and streamlined application of fMRI foundation models.

### Functional connectivity with regularized logistic regression is an important baseline, difficult to beat

Functional connectivity is a low-cost, widely used feature to compute from fMRI data, yet neither foundation model adopted it as an evaluation baseline. Including this baseline links foundation model research to the broader fMRI biomarker literature, where functional connectivity remains the standard feature for trait prediction (Dadi et al., 2019; Finn et al., 2015; Khosla et al., 2019). Our results show that this baseline was hard to beat: functional connectivity outperformed foundation-model features on demographic targets and achieved informative precision on clinical targets, despite requiring no pretraining or architectural changes.

### Limited performance of foundation models on downstream tasks

BrainLM’s original evaluation instead used principal components of the time series as a comparison baseline, reporting that foundation model features outperformed this simpler representation. We constructed the same principal component features for our dataset and found they performed at chance level (see Figure 3), suggesting that the relevant benchmark for fMRI foundation models should be functional connectivity rather than time-series decomposition.

BrainHarmonix’s evaluation, by contrast, benchmarks against several other fMRI foundation models. While this showed improvement over prior foundation-model approaches, the comparison remained limited to that class of methods and did not indicate how these models perform relative to established neuroimaging features such as functional connectivity. Without an external baseline, claims of state-of-the-art performance were difficult to interpret within the broader fMRI biomarker literature. Most importantly, we did not reproduce results in which BrainHarmonix was the definitive winning model compared with BrainLM within foundation models.

### The importance of measuring precision for diagnosis and prognosis

Evaluating machine learning models on clinical datasets presents an additional methodological challenge due to target-class imbalance. Clinical targets are inherently skewed: individuals at a specific disease stage represent a small fraction of the broader sample, and a model that predicts the majority class for every sample achieves high accuracy while failing entirely to identify the cases of clinical interest. Accuracy should therefore never be interpreted without precision in clinical settings (Clarke et al., 2025). Moreover, when a feature or model achieves meaningful precision on a skewed clinical target, this indicates either that the feature carries sufficient variability to differentiate classes or that the model has learned to extract that variability in a clinically useful way, both of which warrant further mechanistic investigation.

When examining precision, BrainLM features exceeded functional connectivity for both prognosis and diagnosis targets, but neither difference was statistically significant. For MCI status (prognosis), the adapted BrainLM attention head CLS token achieved the highest precision but did not reach statistical significance when compared with functional connectivity. This difference is a preliminary pattern, not a reliable effect. However, precision performance was more stable than functional connectivity, given the confidence interval widths. For Aβ status, we saw more foundation-model-derived features outperform functional connectivity, but the well-performing features were inconsistent across the two targets. Notably, neither feature achieved strong discriminative performance overall, as reflected in AUC confidence intervals that included chance level (0.5) for both approaches.

For BrainHarmonix, only features outperformed the functional connectivity baseline in Aβ status prediction, and only slightly exceeded chance in AUC. Yet the top BrainHarmonix performer used no fMRI-derived information. This raised the question of what a T1-weighted structural feature captures about Aβ burden that functional connectivity did not. Yet BrainLM-derived features outperformed BrainHarmonix. It would be interesting to compare these subsets of individuals and investigate the source of differences, such as other clinical, demographic, or imaging characteristics. Identifying what this structural feature captures, why functional connectivity missed it, and when functional neuroimaging data are relevant would be the next step toward understanding whether these models offer genuine added value for early detection of neurodegeneration.

### Limitations and future directions

The current study has a few caveats that future studies should address. Methodologically, we set the hyperparameters using general heuristics. A more extensive hyperparameter search could improve the models and yield features with better predictive performance. The current downtream evaluation workflow was adopted from the source code of BrainLM and optimised for computing resource management, yet it would be important to adopt an end-to-end framework for the downstream evaluation to align the process with standard foundation model research (Thölke et al., 2026). The current downstream tasks are limited to challenging but well-established classification tasks, and the aim is to test the streamlineness of the user adoption experience, rather than seeking optimal performance. These results do not rule out the possibility that, with more extensive changes and improvements, the models could achieve better performance or support more diverse use cases.

Several future research directions could further expand our understanding of the contributions of foundation models to fMRI data and how to improve how we think about fMRI-based foundation models. Investigating the subject profiles of high-precision MCI/Aβ predictions could help us better understand which signals foundation model-based features capture versus noise.

Finally, current studies show that an fMRI foundation model pretrained solely on fMRI signals performs poorly. Two distinct directions warrant investigation. First, Thomas and colleagues pre-trained a self-supervised model on 34 OpenNeuro datasets spanning a deliberately broad range of sites and protocols, rather than a single large cohort, and found that the resulting representations transferred well to new mental state decoding tasks (Thomas et al., 2022); whether this kind of multi-site heterogeneity would improve trait and clinical prediction has yet to be directly addressed. For example, BrainHarmonix includes multisite datasets such as ABCD in the training data, but no assessments were done on state-related prediction. More explicit evaluation is needed. Second, current models are pretrained exclusively on resting-state data, which carries no explicit cognitive context; incorporating naturalistic, cognitively annotated data instead may help models capture representations more directly relevant to the behavioural and cognitive landmarks used in diagnosis. For future directions, a more diverse array of evaluation datasets and targets, including both state- and trait-level evaluation, could indicate whether the latent features of the current foundation model are neurocognitively relevant.

## Conclusion

Taken together, our results clearly indicate that the strong downstream transfer seen in large language models cannot yet be assumed for fMRI foundation models: on this benchmark, neither model is ready for adoption by independent groups without a well-established baseline for comparison and considerable pipeline validation of their own. This highlights the need for standardized libraries for deploying foundation models and for more rigorous evaluation benchmarks featuring high-quality functional connectivity baselines, dummy classifiers, and multiple performance metrics, including precision.

## Data and code availability statement

The dataset is hosted and shared through the Prevent-AD LORIS platform, where we obtained data access (https://preventad.loris.ca). All code for preprocessing, quality assessment metrics, post-processing of the fMRI data, and retrieval of the prediction target is available here: https://github.com/SIMEXP/prevent-ad_dr8.1internal or https://doi.org/10.5281/zenodo.23485573. The code for feature extraction and the main experiments is in a separate repository for simplicity: https://github.com/SIMEXP/prevent-ad-benchmark or https://doi.org/10.5281/zenodo.23485638. The code for results visualization is in https://github.com/SIMEXP/prevent-ad-benchmark-plotting or https://doi.org/10.5281/zenodo.23485584. All second-level analysis results (e.g. model performance scores) and an archive of GitHub repositories are available on Zenodo: <u>10.5281/zenodo.22926041</u>.

## Data Availability

All second-level analysis results (e.g. model performance scores) and an archive of GitHub repositories are available on Zenodo: 10.5281/zenodo.22926041.

https://preventad.loris.ca

## Acknowledgment

The authors wish to acknowledge the PREVENT-AD project for providing the data free of charge, as well as the Brain Imaging Centre of the Douglas Mental Health University Institute. A full listing of the PREVENT-AD University Group members can be found at https://preventad.loris.ca/. We also acknowledge the PREVENT-AD cohort participants for dedicating their time and energy to creating a dataset that benefits the wider community.

## Ethics approval and consent to participate

The PREVENT-AD study protocols, consent forms, and study procedures were approved by the McGill University Institutional Review Board and/or the Douglas Mental Health University Institute Research Ethics Board. All participants provided written informed consent.

## Author contributions (CRediT)

**Hao-Ting Wang**: Conceptualization, Data curation, Formal analysis, Funding acquisition, Methodology, Project administration, Software, Visualization, Writing – original draft, Writing – review & editing

**Alexa Pichet Binette**: Conceptualization, Data curation, Writing – review & editing

**Pravish Sainath**: Methodology, Writing – review & editing

**Nicolas Farrugia**: Methodology, Writing – review & editing

**Sylvia Villeneuve**: Conceptualization, Resources, Writing – review & editing

**Lune Bellec**: Conceptualization, Funding acquisition, Resources, Supervision, Writing – review & editing

